# Vaginal estrogen therapy initiation after breast cancer and oncological outcomes: a nationwide population-based target trial emulation

**DOI:** 10.1101/2024.10.23.24315966

**Authors:** Elise Dumas, Anne-Sophie Hamy, Paul Gougis, Enora Laas, Floriane Jochum, Diane Hill, Christine Rousset-Jablonski, Sophie El Ferjaoui, Philippe-Jean Bousquet, Sophie Houzard, Christine Le Bihan-Benjamin, Fabien Reyal, Kerollos Nashat Wanis, Mats Julius Stensrud, Florence Coussy

## Abstract

**Background:** Genitourinary syndrome of menopause (GSM) frequently occurs after breast cancer (BC), leading to symptoms that can severely affect quality of life. Vaginal estrogen therapies (VETs), including compounds like promestriene and estriol, are recommended for GSMs. However, there is concern that VETs might affect the risk of BC relapse in women with a history of BC.

**Material and Methods:** Utilizing data from the French National Health Data System, we emulated target trials to investigate the effect of VET initiation (promestriene or estriol) on disease-free survival (DFS) in women with a history of non-metastatic BC. Trials were emulated sequentially at 12, 24, 36, and 48 months after BC diagnosis. We estimated survival probabilities at three and five years and used inverse probability weights to adjust for confounders.

**Results:** Of the 136,408 unique patients meeting the inclusion criteria for at least one the emulated trials, 1,737 (1·3%) initiated VET. In patients with hormone receptor (HR)-positive tumors treated with tamoxifen, the estimated difference in DFS for VET initiation *versus* no initiation was 0·2 percentage-point at five years (95% CI −4·1; 3·6), while it was −2·9 (95% CI −6·5; 0·0) in patients with HR-positive tumors treated with aromatase inhibitors. In this subgroup, the estimated difference in DFS for promestriene initiation *versus* no VET initiation was −2·5 percentage-points at five years (95% CI −6·7; 1·1) while it was −3·7 (95% CI −10·1; 1·8) for estriol initiation *versus* no VET initiation; and the differences between the two molecules were even more pronounced at three years.

**Discussion:** Our results do not find evidence that VET decreases DFS in patients with *HR*-positive tumor treated with tamoxifen. However, VET initiation might decrease DFS in patients treated with aromatase inhibitors, with estriol leading to a more pronounced decrease in DFS than promestriene.

## Introduction

Breast cancer (BC) is the most frequent cancer in women, with approximately 2·3 million cases reported annually [1]. Due to the high incidence of BC around menopausal age and the common use of endocrine therapy as adjuvant treatment, BC survivors often experience symptoms associated with declining estrogen levels [2], including genitourinary syndrome of menopause (GSM). GSM is primarily characterized by vaginal dryness, itchiness, burning, overactive bladder, urinary incontinence or infection, sexual symptoms, and significantly impaired quality of life [3].

Vaginal estrogen therapies (VETs) are currently the main recommended treatment for the management of GSMs [4,5]. Available molecules include conjugated equine estrogens, estradiol (E2), estriol (E3), the final metabolite of estrogen synthesis, and promestriene, a synthetic estrogen-based VET. While all of these formulations are approved as pharmacologic interventions for the treatment of isolated moderate-to-severe GSM in healthy women [6,7], their availability varies worldwide. For example, promestriene is available in some European countries, such as France, but not in the United States. In case of history of BC, both the U.S. Food and Drug Administration and European Medicines Agency formally contraindicate the use of VET. However, most national and international guidelines, albeit cautious, endorse the use of VET after BC as a second-line intervention in case of non-hormonal treatments failure, with shared decision making between the oncologist and the patient [7–10].

Systemic administration of estrogen can lead to BC relapse [11]. Indeed, the rationale for prescribing endocrine therapy (ET) is to block estrogen pathways, either by reducing estrogen synthesis or by competitive linkage with estradiol receptors. However, current literature evaluating the effect of topical VET on oncological outcomes in patients with previous BC has yielded inconsistent results. The reported effect of VETs on serum estradiol levels varied according to studies, with some studies finding increased estradiol levels[12,13] while others did not find changes [14–16]. From an epidemiological view, most observational studies found no significant association, or a positive association, between VET use after BC and BC prognosis [17–25]. However, two studies found an increased risk of relapse in the subgroup of patients treated with aromatase inhibitors (AIs) in patients using VETs [24,25]. So far, no randomized controlled trial has been conducted to evaluate the safety of VET after BC with recurrence as a primary endpoint, leaving open the question of the safety of VET utilization after BC.

Previous observational studies relied on classical statistical methods and were not designed to answer causal questions. In contrast, target trial emulation has been recently developed and popularized as a framework for designing observational studies that aim to quantify effects of interventions [26,27]. This approach consists of designing the observational study analysis to mimic key features of a hypothetical target randomized controlled trial in which the interventions would be compared, thereby reducing the risk of fundamental design flaws and highlighting potential sources of bias that could result in spurious causal conclusions.

In this study, we used data derived from one of the largest population-based resources available so far - the French National Health Data System [28,29] - to emulate target trials investigating the effect of VET initiation during BC follow-up on disease-free survival (DFS).

## Material and methods

### Data source, ethics, and data protection

We used nationwide data from the previously published retrospective French Early Breast Cancer Cohort (FRESH) cohort [30], which was released from the French administrative health care database, SNDS (Système National des Données de Santé) within the Oncology Data Platform available at the French National Cancer Institute [28,29]. The FRESH cohort includes all women 18 years and older, with non-metastatic BC newly diagnosed between January 1, 2011, and December 31, 2017. Details about the cohort constitution and the data available have been previously published [30] and are provided in the Supplementary Notes. The study was conducted in collaboration between Institut Curie and the French National Cancer Institute (INCa), following institutional and ethical rules concerning research involving patient data, and was authorized by the French data protection agency (Commission nationale de l’informatique et des libertés—CNIL) under registration number 920017. No informed consent was required because the data used in the study was de-identified.

### Hypothetical target trial design

We begin by defining a hypothetical target randomized controlled trial that would allow us to identify effects of VETs on relevant clinical outcomes. Inclusion criteria for the hypothetical target trial were (i) being a woman with a history of non-metastatic BC, treated at least by breast surgery and any systemic treatment (chemotherapy, targeted therapy, or endocrine therapy), (ii) being 70 years of age or younger, (iii) not having a history of BC recurrence, and (iv) not having used VET within the 2 years before target trial inclusion. In the hypothetical target trial, individuals were randomly assigned to one of two arms: (i) initiation of VET (treatment arm), or (ii) no initiation of VET (control arm). The trial endpoint was disease-free survival (DFS), defined as the time, in months, from BC diagnosis to death, loco-regional recurrence, contralateral recurrence or distant recurrence, whichever occurred first. The causal estimand of interest was the average treatment effect (ATE) of initiating VET, which could be estimated by the difference in DFS probabilities in the two arms at three years and five years after trial inclusion. Subgroup analyses with respect to hormone receptor (HR) status and type of endocrine therapy (ET) were conducted.

### Emulation using observational data

We next describe how the hypothetical target trial was emulated using the SNDS data. The emulation step involves matching the conditions specified in the hypothetical target trial as closely as possible.

#### Inclusion criteria

Patients were included in the emulated trial if they met the inclusion criteria of the hypothetical target trial and had not yet been administratively censored. As a single patient may meet the eligibility criteria on multiple occasions, we emulated not a single trial, but four trials sequentially, starting at months 12, 24, 36, and 48 after BC diagnosis, respectively. A patient could be included in several of these emulated trials, as long as she met the inclusion criteria on the month of trial onset.

#### Treatment assignment

For each of the four emulated trials, patients meeting the inclusion criteria were classified as initiating VET if they redeemed at least two prescriptions of VET in the 12 months after inclusion, and before relapse or death. The 12-month period during which patients were permitted to initiate VET is referred to as the grace period. VET delivery was identified based on the outpatient delivery of estriol or promestriene, presented either as cream or vaginal ovules (Supplementary Table 1). The control group comprised patients who were not delivered VET in the year after inclusion in the emulated trial. Patients with a single VET delivery were considered to have neither initiated nor refrained from having initiated the treatment and were not included in either arm of the emulated trials.

#### Oncological outcome

The oncological outcome was disease free survival (DFS), as defined in the hypothetical trial. Relapses were identified from the treatments received and the conditions recorded during hospitalizations stays (Supplementary Notes and Supplementary Table 2). All patients were administratively censored on March 1, 2019, and administrative censoring was assumed to be non-informative.

#### Covariates

Covariates could either be measured once, at the time of BC diagnosis and initial treatment, in which case they were fixed for a single patient across all emulated trials, or they could be measured repeatedly at the time of inclusion for each of the emulated trials, in which case they could vary for a single patient across all emulated trials. Covariates measured once included the (1) number of general practitioner visits, (2) number of gynecologist visits, and (3) performance of mammography screening in the year before BC diagnosis; (4) deprivation level of the area of residence and (5) enrollment in universal health insurance at BC diagnosis; (6) BC subtype, (7) nodal status, (8) type of BC surgery, (9) use of axillary surgery, (10) type of hospital for surgery, (11) use of radiotherapy and (12) chemotherapy setting during initial BC treatment. Covariates measured repeatedly included (1) endocrine therapy regimen, (2) adherence to endocrine therapy, (3) comorbid conditions, (4) exposure to other medications at trial inclusion, and (5) age at trial inclusion. Further details on the definition of covariates are provided in the Supplementary Notes.

#### Statistical analysis

Unlike the hypothetical target trial, treatment assignment in the observed data was not randomized and could depend on a patient’s characteristics, potentially leading to confounding. Without appropriate adjustment, the study would also be subject to immortal-time bias due to the 12-month grace period allowed between inclusion and VET initiation. To address both confounding and immortal-time bias, we cloned each trial participant into two exact copies (clones) [31,32]. One clone was assigned to the treatment arm, the other to the control arm. Clones were artificially censored when their observed trajectories deviated from the treatment strategy of the arm to which they were assigned. This informative artificial censoring introduces selection bias over time, which was addressed by the use of time-varying inverse probability of censoring (IPC) weights, estimated from the covariates. Standardized mean differences (SMDs) were derived to assess adjustment quality after IPC weighting.

The ATE of VET initiation was estimated by the three-year and five-year differences in DFS on the IPC-weighted population and was visually assessed by weighted Kaplan-Meier survival curves. We intentionally reported outcomes on the survival scale to avoid the problems associated with the causal interpretation of hazard ratios [31,33]. We conducted subgroup analyses by HR status (*HR*-positive/*HR*-negative) and endocrine therapy regimen (tamoxifen/aromatase inhibitors), and we analyzed the two molecules used as VET (estriol/promestriene) both combined and separately. We performed a sensitivity analysis to assess potential residual confounding bias, using fosfomycin, a first-line antibiotic used in the treatment of urinary tract infections (UTIs) as a negative control exposure [34]. Fosfomycin emerged as a suitable candidate for a negative control, because vaginal dryness and UTIs are both GSMs that could be induced by hypoestrogenic state [35], suggesting a common confounding structure of VET and fosfomycin, and because there was no prior evidence that fosfomycin could affect BC outcomes.

In a supplementary analysis, we adopted a methodology similar to previously proposed approaches [23], utilizing a time-dependent Cox proportional hazards regression model to estimate hazard ratios (HRs). Further details are provided in the Supplementary Notes and in Supplementary Table 3.

## Results

### Patients’ characteristics

Overall, 136,408 patients with previous BC met the inclusion criteria for at least one of the emulated trials, representing 373,568 patient-years in the aggregated analysis (Supplementary Figure 1). Of them, 1,737 patients (1·3%), representing 1,739 patient-years, initiated VET (Supplementary Table 4). Most patient-years who initiated VET received only promestriene (55·5%), while 34·6% received only estriol, and 9·9% received both promestriene and estriol (Supplementary Figure 2A). The median number of VET deliveries was 3 (IQR 2-6, Supplementary Figure 2B).

When compared with patients who did not initiate VET, patients who initiated VET were slightly older at BC diagnosis (mean age 54·9 *versus* 54·0), resided in less deprived areas and had more frequent visits to their general practitioner and gynecologist during the year preceding BC diagnosis (Table 1). Overall, BC subtype was mostly luminal (77·3%) while TNBC (10·9%) and *HER2*-positive BC subtypes (*HR*-positive 7·6%; *HR*-negative 4·2%) were less frequent, and this distribution was not different between patients who initiated VET and patients who did not initiate VET. Regarding covariates measured repeatedly, patient-years who initiated VET were more frequently treated with AI at trial inclusion (65·0% *versus* 57·9% of patients with *HR-*positive tumors) while less frequently treated with tamoxifen (Supplementary Table 5).

**Table 1:** Patient’s characteristics by VET initiation status for all variables measured at the time of BC diagnosis and initial treatments. Patients were assigned to the “Any VET” group if they were classified as patients initiating VET in at least one of the four emulated trials; otherwise, they were assigned to the “No VET” group. The number of patients, and the percentage of patients (in parentheses), are reported for categorical variables. The mean value, and the standard deviation (in parentheses), are reported for continuous variables. Of note: of the 136,408 patients meeting the inclusion criteria for at least one of the emulated trials, 1,466 patients received VET only once during the grace period for at least one of the emulated trials, and were artificially censored from both arms for this trial. They are not shown in this table for the sake of simplicity. *Number of general practitioner (GP) visits in the year preceding BC diagnosis. **Number of gynecologist visits in the year preceding BC diagnosis. Abbreviations: VET: vaginal estrogen therapy; GP: general practitioner; BC: breast cancer; TNBC: triple-negative breast cancer; CLCC: “Centre de Lutte Contre le Cancer” (center for the fight against cancer); HR: hormone receptor; HER2: human epidermal growth factor receptor 2.

|  |  | No VET | Any VET |
| --- | --- | --- | --- |
| *Number of patients* |  | 133205 | 1737 |
| Socio-economic factors |  |  |  |
| Age at BC diagnosis |  | 54.0 (9.5) | 54.9 (8.4) |
| Deprivation index | 1st quintile (least deprived) | 25099 (18.8) | 414 (23.8) |
|  | 2nd quintile | 26457 (19.9) | 340 (19.6) |
|  | 3rd quintile | 26020 (19.5) | 324 (18.7) |
|  | 4th quintile | 26365 (19.8) | 320 (18.4) |
|  | 5th quintile (most deprived) | 26637 (20.0) | 298 (17.2) |
|  | Overseas departments | 2627 (2.0) | 41 (2.4) |
| GP visits* | 0 | 8688 (6.5) | 75 (4.3) |
|  | 1-5 | 69124 (51.9) | 812 (46.7) |
|  | 6-11 | 40699 (30.6) | 574 (33.0) |
|  | 12+ | 14694 (11.0) | 276 (15.9) |
| Gynecologist visits** | 0 | 67235 (50.5) | 733 (42.2) |
|  | 1 | 39471 (29.6) | 536 (30.9) |
|  | 2-3 | 22138 (16.6) | 378 (21.8) |
|  | 4+ | 4361 (3.3) | 90 (5.2) |
| Enrollment in universal health insurance | No | 125157 (94.0) | 1657 (95.4) |
|  | Yes | 8048 (6.0) | 80 (4.6) |
| BC diagnostic, biology, and treatment |  |  |  |
| Mammographic screening before BC diagnosis | No | 81645 (61.3) | 1018 (58.6) |
|  | Yes | 51560 (38.7) | 719 (41.4) |
| BC subtype | luminal | 102977 (77.3) | 1367 (78.7) |
|  | TNBC | 14492 (10.9) | 181 (10.4) |
|  | HER2+/HR+ | 10176 (7.6) | 127 (7.3) |
|  | HER2+/HR- | 5560 (4.2) | 62 (3.6) |
| HR status | HR -positive | 113153 (84.9) | 1494 (86.0) |
|  | HR -negative | 20052 (15.1) | 243 (14.0) |
| Nodal status | Node-negative | 102328 (76.8) | 1389 (80.0) |
|  | Node-positive | 30877 (23.2) | 348 (20.0) |
| Type of BC surgery | Lumpectomy | 98061 (73.6) | 1354 (78.0) |
|  | Mastectomy | 35144 (26.4) | 383 (22.0) |
| Axillar surgery | No | 10201 (7.7) | 155 (8.9) |
|  | Yes | 123004 (92.3) | 1582 (91.1) |
| Type of hospital for surgery | CLCC | 36537 (27.4) | 480 (27.6) |
|  | Non profit private hospitals | 4639 (3.5) | 54 (3.1) |
|  | Profit private hospitals | 55408 (41.6) | 776 (44.7) |
|  | Public hospitals | 36621 (27.5) | 427 (24.6) |
| Radiotherapy | No | 10019 (7.5) | 122 (7.0) |
|  | Yes | 123186 (92.5) | 1615 (93.0) |
| Chemotherapy | No | 58983 (44.3) | 806 (46.4) |
|  | Yes | 74222 (55.7) | 931 (53.6) |
| Setting | Adjuvant | 59074 (79.6) | 762 (81.8) |
|  | Neo-adjuvant | 13678 (18.4) | 148 (15.9) |
|  | Both | 1470 (2.0) | 21 (2.3) |

After adjustment by IPC weighting, all SMDs were below 0·1 in absolute value (Supplementary Figure 3), suggesting that the statistical procedure enabled effective balancing of the characteristics between the group initiating VET and not initiating it. The estimated IPC weights at the end of the 12-month grace period ranged from 1·28 to 3·29 (median 2·04, interquartile range IQR 1·84-2·31, Supplementary Figure 4). The median follow-up was 33 months (IQR: 16-50).

### Effect of VET initiation on DFS

In the whole population, the estimated difference in DFS of VET initiation *versus* no initiation was 0·3 percentage-point at three years (95% CI −1·1 to 1·4) and −1·5 at five years (95% CI - 3·7 to 0·6) (Figure 1A, Table 2). In patients with HR-positive tumors, the estimated difference at five years was −1·9 (95% CI −4·6 to 0·3, Figure 1B). We further subcategorized patients with HR-positive tumors by ET regimen. In patients treated with AIs, the estimated difference in DFS was −0·6 percentage-point at three years (95% CI −2·6 to 0·9), and −2·9 at five years (95% CI −6·5 to 0·0) (Figure 1C). Conversely, in patients treated with tamoxifen, the estimated difference in DFS was 2·2 percentage-points at three years (95% CI 0·4 to 3·7) and 0·2 at five years (95% CI −4·1 to 3·6, Figure 1D). In patients with *HR*-negative tumors, the estimated difference was 0·3 percentage-point at three years (95% CI −3·4 to 3·9) and 1·9 at five years (95% CI −2·4 to 6·7) (Figure 1E).

**Figure 1:**
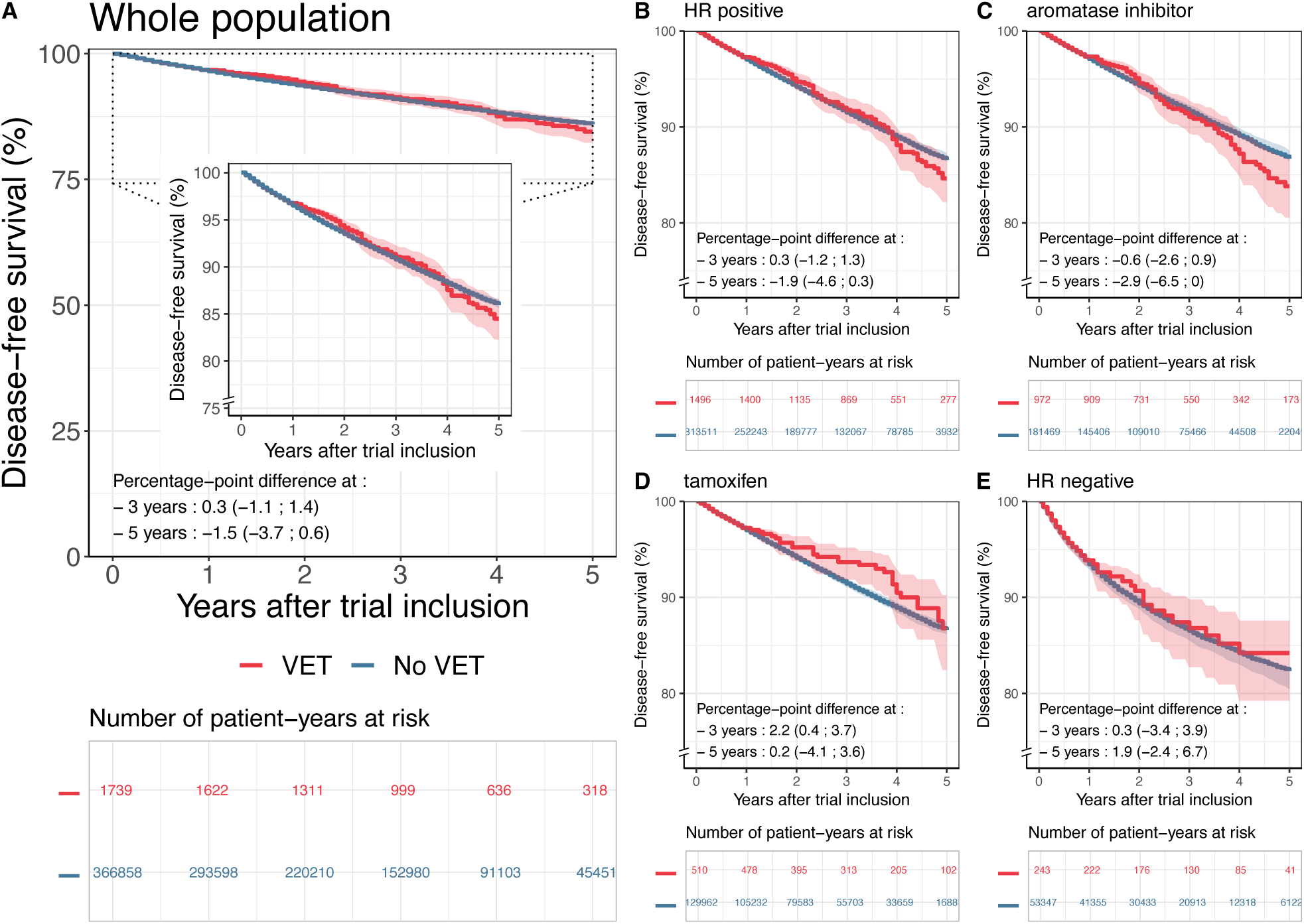
Effects of VET initiation on disease-free survival in the whole population of BC patients (A) and for subgroups defined by HR status and endocrine therapy regimen (B-E). Abbreviations: VET: vaginal estrogen therapy; HR: hormone receptor; CI: confidence interval.

**Table 2:**
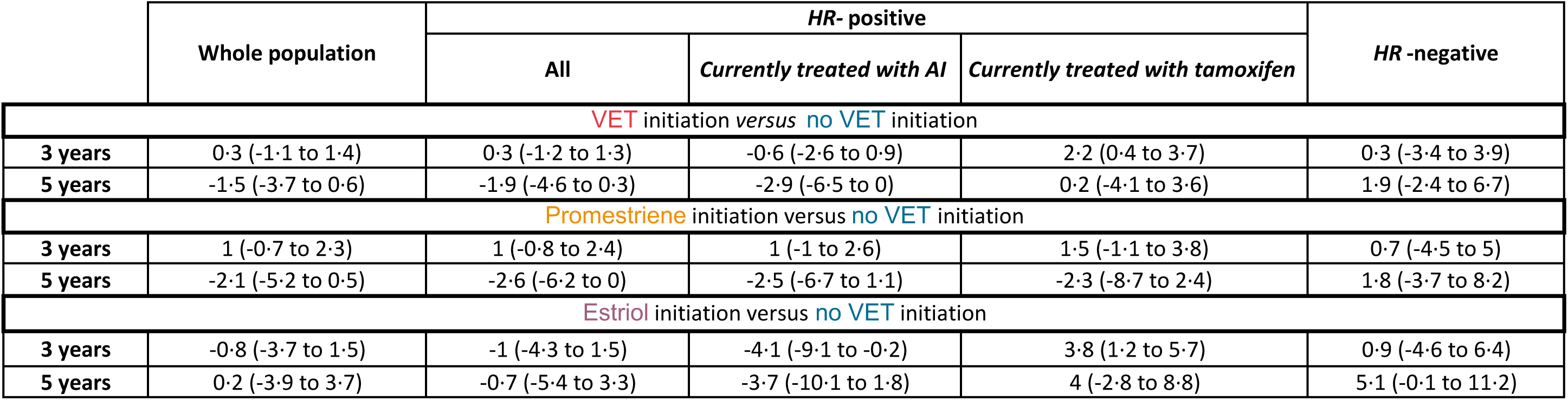
Difference in disease-free survival (DFS, in percentage-points) at three years and five years and associated 95% confidence intervals for VET initiation *versus* no VET initiation, promestriene initiation *versus* no VET initiation, and estriol initiation *versus* no VET initiation for the whole population and per subgroup. Abbreviations: VET: vaginal estrogen therapy; HR: hormone receptor; AI: aromatase inhibitor.

### Effect of VET initiation on DFS according to the VET molecule

The estimated differences in DFS of promestriene initiation *versus* no VET initiation are displayed in Figure 2. In patients with HR-positive tumors treated with AI, the estimated difference in DFS was 1·0 percentage-point at three years (95% CI −1·0 to 2·6) and −2·5 percentage-points at five years (95% CI −6·7 to 1·1) (Figure 2C, Table 2).

**Figure 2:**
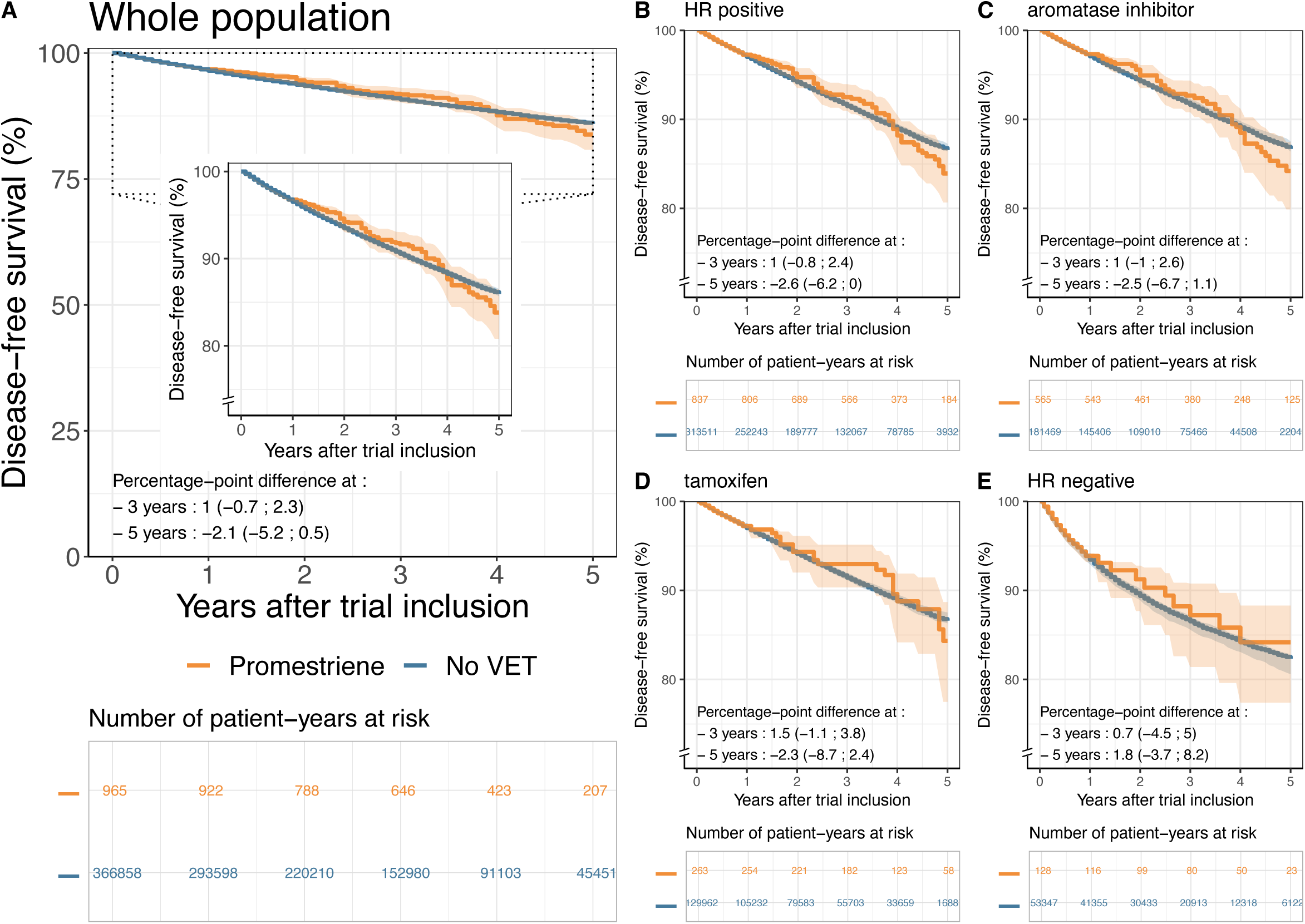
Effects of promestriene initiation on disease-free survival in the whole population of BC patients (A) and for subgroups defined by HR status and endocrine therapy regimen (B-E). Abbreviations: VET: vaginal estrogen therapy; HR: hormone receptor; CI: confidence interval.

The estimated differences in DFS of estriol initiation *versus* no VET initiation are displayed in Figure 3. Among patients with HR-positive disease currently treated with AIs, the estimated difference in DFS was −4·1 percentage-points at three years (95% CI −9·1 to −0·2) and −3·7 at five years (95% CI −10·1 to 1·8) (Figure 3C); while it was 3·8 percentage-points at three years (95% CI 1·2 to 5·7) and 4·0 at five years (95% CI −2·8 to 8·8) among patients currently treated with tamoxifen (Figure 3D).

**Figure 3:**
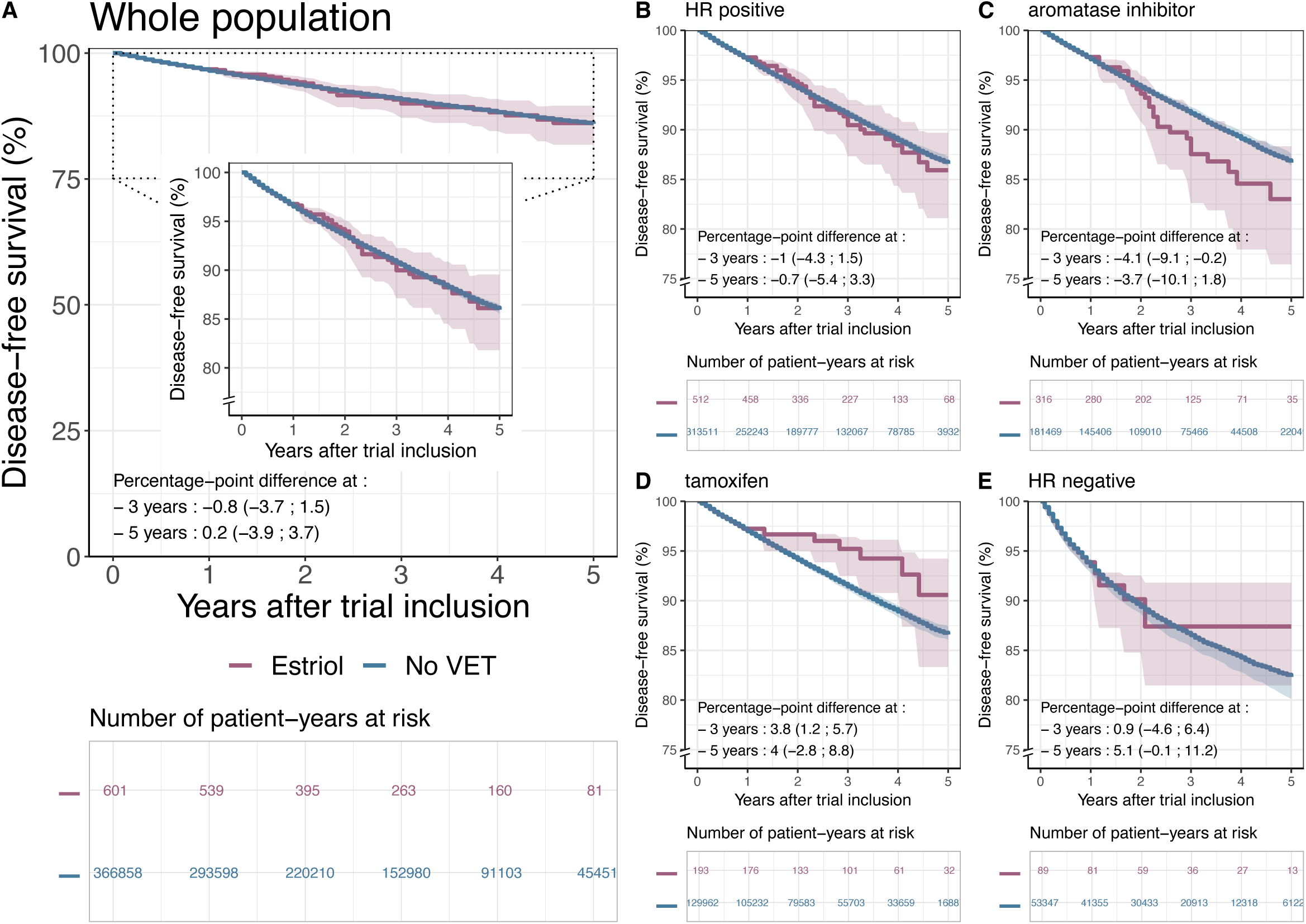
Effects of estriol initiation on disease-free survival in the whole population of BC patients (A) and for subgroups defined by HR status and endocrine therapy regimen (B-E). Abbreviations: VET: vaginal estrogen therapy; HR: hormone receptor; CI: confidence interval.

### Negative control exposure

We analyzed fosfomycin initiation as a negative control exposure. The estimated difference in DFS for fosfomycin initiation *versus* no initiation was low in the whole population, and in patients with *HR*-positive tumors, regardless of their endocrine therapy regimen, both at three and five years (Supplementary Tables 6-7, Supplementary Figure 5). However, the estimated difference in DFS was 4·7 percentage-points (95% CI 0·8 to 7·8) at five years in patients with *HR*-negative tumors (Supplementary Figure 5E).

### Comparison with a time-dependent Cox model

In a supplementary analysis mimicking an associational method previously proposed,[23] the estimated HR of VET initiation *versus* no initiation was 0·95 (95% CI 0·89-1·00) in the whole population, 0·96 (95% CI 0·82-1·12) in the subgroup of patients treated with tamoxifen, and 0·93 (95% CI 0·87-1·00) in the subgroup of patients treated with aromatase inhibitors (Supplementary Table 3).

## Discussion

We emulated target trials investigating the effect of VET initiation on the risk of relapse or death in patients with non-metastatic BC. We found an increased risk of relapse of 2·9 percentage-points at five years among patients initiating VET while treated by AI, notably in patients who initiated estriol.

Previous studies on VET use after BC provided reassuring results (Supplementary Table 8). None found an adverse association between the use of VET and overall survival (OS) [20,21,25], BC-specific survival [20–23], or DFS [17–21,25] in the broad population of BC survivors. On the contrary, some reported that VET use is associated with improved OS [25], BC-specific survival [21,23], or DFS [21]. In particular, in the largest study so far, McVicker et al [23] analyzed BC-specific survival according to VET use in a cohort of 49,237 women with BC aged between 40 and 79 years. The authors found that VET use was associated with a decrease in specific mortality (HR 0·77, 95% CI 0·63 to 0·94) in the overall population, but also in the subgroup of patients treated with AIs (HR 0·72, 95% CI 0·58 to 0·91). In contrast, a Danish observational study reported VET use to be associated with an increased risk of recurrence in this subgroup (HR 1·39, 95% CI 1·04 to 1·85) [25].

All the previous observational studies are difficult to interpret causally and are prone to bias because they did not apply formal causal inference methods. In our work, by explicitly emulating a target trial, we have a straightforward causal interpretation of our results under explicit assumptions. In contrast, earlier investigations relied on traditional associational methods, estimating HRs from time-fixed or time-varying Cox models in cohort designs [18,20,21,23–25], or odds ratios (ORs) from logistic regression models in case-control designs [17,22]. These methods do not provide valid causal estimates, as shown in our supplementary analysis where, by mimicking the methodology used in a previous study [23], we estimated the HR in the AI group to be 0·93 (95% CI 0·87-1·00), in the opposite direction to the results we obtained in our main analysis. Several reasons may explain this difference [32,36]. First, traditional methods cannot account for treatment-confounding feedback due to time-varying confounders [37–40]. In contrast, in our main analysis, we handled time-varying confounding factors by emulating multiple target trials sequentially, treating time-varying confounding factors as time-fixed confounders at each trials’ baseline. Second, HRs and ORs are not collapsible, which implies that the covariate-adjusted HRs from the previous observational studies are substantially different quantities than the HRs estimated in randomized clinical trials [33]. More broadly, there are concerns about the causal interpretation of HRs [31]. This justifies why we have only presented results at the survival scale. Third, traditional time-fixed methods are susceptible to immortal-time bias, arising when a grace period for treatment initiation leads to a spurious overestimation of the protective effect of treatment [41]. We avoided the bias by a cloning and weighting approach. Finally, any observational study might be prone to bias due to unmeasured confounding. Unlike previous studies, we used a negative control exposure, fosfomycin, to detect potential residual confounding bias. The negative control analysis did not detect evidence of unmeasured confounding in any subgroup, except *HR*-negative tumors. Thus, we warrant caution in interpreting results pertaining to the *HR*-negative group. Conversely, the results of the negative control exposure analysis support a causal interpretation of our findings in the remaining subgroups.

Overall, in the current study, no harmful effect of VET was detected marginally, in the overall population. However, among patients with HR-positive tumors, the effect of VET initiation on DFS was different according to ET regimen, with a transient protective effect of VET initiation in patients under tamoxifen at three years, not confirmed at five years; and a deleterious effect of VET initiation in patients under AI at five years. Several randomized trials have reported transient increases in circulating estrogen levels associated with the use of VET [12,42], with systemic passage potentially exacerbated by vaginal atrophy [16]. Tamoxifen, a selective estrogen receptor modulator, competes with estrogens and their derivatives with a higher affinity for the estrogen-receptor, resulting in the inhibition of estrogen oncogenic pathway, thereby counteracting the effects of small increases in circulating estrogen levels [43]. The increase in DFS at three years observed in patients initiating VET while on tamoxifen could be due to improved adherence to endocrine therapy, but further research is needed to validate this hypothesis. Conversely, AIs inhibit estrogen production to nearly undetectable circulating levels by inhibiting the conversion of adrenal androgens to estrogens [44]. With results suggesting that long-term sharp estrogen deprivation may cause a hypersensitivity of estrogen receptors in the breast [16], even a modest increase in estrogen levels in patients treated with AIs could promote cancer recurrence.

Among AI-treated patients, we observed decrease in DFS in patients initiating estriol at three years, but not in patients initiating promestriene. Several previous studies suggested that the systemic passage of VETs may vary by molecule, with promestriene exhibiting limited systemic absorption [45]. Promestriene has a favorable safety profile with no alteration of plasmatic levels of gonadotrophins or estradiol [46,47] while variations were found with estriol [14,48], no stimulation of endometrium [49], and a pharmacological specificity with the presence of 3-propyl and 17b-methyl ether groups making it less able to penetrate the basal membrane.

We report here the largest study from far on the effect of VET initiation on oncological outcomes. While we used state-of-the-art causal inference methodology, our study has limitations. Firstly, our data do not allow assessment of over-the-counter purchases such as non-hormonal vaginal moisturizers, estradiol vaginal rings or prasterone. Secondly, we were not able to provide data on patients currently treated with gonadotropin-releasing hormone (gnRH) agonists together with AI or tamoxifen, due to the timeframe of the study, where these combinations were not yet standard of care. Finally, we were unable to draw conclusions for the subgroup of patients with *HR*-negative tumors due to potential residual unmeasured confounding, highlighting an area for future research.

In conclusion, our study suggests that VET initiation during BC follow-up could be safely proposed to patients with HR-positive tumors currently treated with tamoxifen. In patients currently treated with AIs, VET initiation should be avoided as much as possible until mature additional data are available. In the absence or after failure of non-hormonal alternatives, promestriene should be preferred over estriol in this subgroup. In a context of a growing number of patients are treated under AI notably with gnRH agonists, the development of novel efficient non hormonal therapies to alleviate GSM remains a burning unmet need.

## Data Availability

All data produced in the present study are available upon reasonable request to the authors.

## Acknowledgments

We thank the Department of Health Data and Assessment, Health Survey Data Science and Assessment Division, French National Cancer Institute (Institut National du Cancer INCa) for providing us with access to the cancer cohort.

## Funding

This study was funded by Monoprix*, INCa grant number 18-127 within the COMBIMMUNO (Comedications and comorbidities in breast cancer: Deciphering Interactions Between Immune Infiltration, Response to Treatment and Prognosis) project, and the Swiss National Science Foundation. The funder was not involved in study design, or in the collection, analysis, and interpretation of data, the writing of this article or the decision to submit it for publication.

## Competing interests

Christine Rousset-Jablonski reports payment or honoraria for lectures, presentations, speakers bureaus, manuscript writing or educational events by Theramex (payment made to institution). Paul Gougis reports consulting fees for BMS, academic grant from Sanofi and travel accommodation by Eisai. Other authors have no relevant financial or non-financial interests to disclose.

## Author Contributions

(I) Project Conception: Ame-Sophie Hamy, Fabien Reyal, Florence Coussy.
(II) Statistical Study design: Kerollos Nashat Wanis, Mats Julius Stensrud, Elise Dumas.
(III) Data Provision and quality: Philippe-Jean Bousquet, Sophie Houzard, Christine Le Bihan-Benjamin.
(IV) Medical support : Enora Laas, Floriane Jochum, Diane Hill, Christine Rousset-Jablonski, Sophie El Ferjaoui, Paul Gougis, Florence Coussy.
(V) Data analysis and interpretation: Elise Dumas.
(VI) Manuscript first draft: Elise Dumas, Mats Stensrud, Kerollos Wanis, Anne-Sophie Hamy, Florence Coussy.
(VII) Manuscript writing: All authors.
(VIII) Final approval of manuscript: All authors.

## Ethics approval

The study was conducted in collaboration between Institut Curie and the French National Cancer Institute (INCa), following institutional and ethical rules concerning research involving patient data, and was authorized by the French data protection agency (Commission nationale de l’informatique et des libertés—CNIL) under registration number 920017.

## Consent to participate

In accordance with French regulations applicable to the SNDS, no informed consent was required because the data used in the study was de-identified.

## Notes

### Author Declarations

Ethics committee of French data protection agency (Commission nationale de l informatique et des libertes, CNIL) gave ethical approval for this work

