## Supplementary Material for "Vaginal estrogen therapy initiation after breast cancer and oncological outcomes: a nationwide population-based target trial emulation"

### Material and methods

#### Data source, ethics and data protection

The FRESH database includes the SNDS data of all patients included in the FRESH cohort, which were identified with the Oncology Data Platform (ODP) available at the French National Cancer Institute (INCa). SNDS data has been described in detail elsewhere.[1,2] Briefly, it includes (i) demographic data (sex, date of birth, zip code of the town of residence, vital status, date of death if appropriate, health insurance regimen), (ii) hospital discharge reports (diagnoses, medical procedures, and expensive treatments), (iii) outpatient care (drugs dispensed, with the date of delivery, laboratory tests, and outpatient medical procedures) and (iv) long-term illness (LTI) records. Demographic data, hospital discharge reports and outpatient care data were available for the year preceding patient inclusion, up to December 31, 2018. Data for the patients' history of LTI were available until December 31, 2018. The diagnosis codes were recorded in the SNDS, based on the International Classification of Diseases – 10th revision, ICD-10.[3] Procedures were recorded with the CCAM classification (*Classification Communes des Actes Médicaux*). Medications prescribed in outpatient care were recorded with CIP (*Code Identifiant de Présentation*) codes. In hospital, only costly innovative drugs included in a special reimbursement process called "*list en sus*" were recorded, in the form of UCD (*Unités Communes de Dispensation*) codes. Both the UCD and CIP codes were linked to the ATC classification (Anatomical Therapeutic and Chemical classification) of the World Health Organization. Medical devices reimbursed by the French health insurance system (external prosthetics, orthotics, active implantable medical devices, invalid carriages, medical beds etc.) were recorded with LPP (*Liste des produits et prestations*) codes. In outpatient care, the type of medical service (teleconsultation, nursing care, dental care, etc.) was recorded with NGAP (*Nomemenclature Générale des Actes Professionnels*) codes and the physician's specialty was recorded with an untitled nomenclature (variable *PSE_SPE_COD*).

#### Hypothetical target trial design

In the hypothetical target trial, the choice of the specific type of drug and presentation would have been left to the physician, with combinations allowed. Both patients and their healthcare teams would be aware of the patient's treatment status. Consistency and unbiasedness of the difference in survival probabilities three and five years after trial inclusion as an estimator of the average treatment effect (ATE) requires no informative loss to follow-up, consistency, ignorability, and positivity.[4] Ignorability and positivity would have been ensured by experimental design.[5]

#### Emulation using observational data

##### Treatment assignment

VET intake was identified based on the outpatient delivery of vaginal estrogen formulations under prescription (Supplementary Table 1). We considered patients with a single VET delivery to be at high risk of not adhering to treatment, so we did not include them as patient who initiated VET. They were also not included in the reference group.

##### Outcome

The occurrence of any of loco-regional recurrence, distant recurrence, or contralateral recurrence, was identified based on (i) the resumption of radiotherapy, chemotherapy, or targeted therapy at least six months after the end of the initial treatments, (ii) a breast surgery procedure with axillar procedure performed at least six months after the end of the initial treatments, (iii) the intake of an anti-cancer molecule approved only in the metastatic setting starting at least six months after initial breast surgery, or (iv) the presence of a diagnosis code of metastasis in hospitalization stays starting at least six months after initial breast surgery. We defined the date of BC relapse as the first day of any of the above events. The list of anti-cancer molecule approved only in the metastatic setting and the list of diagnosis code of metastasis is available in Supplementary Table 2.

##### Covariates

1) *Number of general practitioner (GP) visits in the year preceding BC diagnosis:* General practitioner (GP) visits were identified by outpatient care visits with specialist code “1” (general practice), “22” (general practice specialist with diploma) or “23” (general practice specialist acknowledged by the French Medical Board). The *number of GP visits in the year preceding BC diagnosis* was calculated as the number of days with at least one GP visit during the 365 days preceding BC diagnosis. This variable was binned into the following categories: (1) 0 (no GP visits in the year preceding BC diagnosis); (2) 1-5 (one to five visits in the year preceding BC diagnosis); (3) 6-11 (six to eleven visits in the year preceding BC diagnosis); (4) 12+ (twelve or more visits in the year preceding BC diagnosis).
2) *Number of gynecologist visits in the year preceding BC diagnosis:* Gynecologist visits were identified by outpatient care visits with specialist code “7” (obstetric gynecologist), “70” (medical gynecologist), “77” (obstetrician) or “79” (obstetric and medical gynecologist). The *number of gynecologist visits in the year preceding BC diagnosis* was calculated as the number of days with at least one gynecologist visit among the 365 days preceding BC diagnosis. The variable was binned into the following categories: (1) 0 (no gynecologist visit in the year preceding BC diagnosis); (2) 1 (one visit in the year preceding BC diagnosis); (3) 2-3 (two or three visits in the year preceding BC diagnosis); (4) 4+ (four or more visits in the year preceding BC diagnosis).

3) *Performance of a mammography screening in the year preceding BC diagnosis:* screening mammograms were identified based on the presence of the procedure code QEQK004 in both hospital and outpatient care records in the year preceding breast cancer diagnosis. The variable was binned into the following categories: 'yes' if the patient underwent a screening mammography in the year preceding breast cancer diagnosis, or 'no' otherwise.
4) *Deprivation level:* The area of residence was defined as the zip code of the French '*département*' (equivalent to a county) of residence at the time of first BC surgery. We used the 'FDep09' geographic socioeconomic index as a measure of social deprivation, as described elsewhere.[6] This index was defined at the '*commune*' (the smallest administrative unit in France) level in 2009, exclusively for mainland France. The *patient's deprivation index* was set as the mean 'FDep09' index for the '*communes*' included in the patient's *département* of residence in mainland France. It was set as 'missing' for overseas *départements*. The *deprivation index* was classified into six categories: (1) "Overseas *départements*"; and the five quintiles of the distribution for patients living in mainland France: (2) "1st quintile (least deprived)", (3) "2nd quintile", (4) "3rd quintile", (5) "4th quintile" and (6) 5th quintile (most deprived).
5) *Enrollment in universal health insurance at BC diagnosis*: Patients were considered enrolled in universal health insurance if they were covered by the "*Couverture Maladie Universelle Complémentaire (CMU-C)*" at the time of BC diagnosis. CMU-C is a government-sponsored complementary universal public insurance scheme in France, proposed for low-income individuals. The information was directly available in the SNDS database.
6) *BC subtype*: BC subtype was derived from the BC treatments received,[7] and categorized into (1) luminal, (2) TNBC (triple-negative breast cancer), (3) HER2-positive (human epidermal growth factor receptor 2-positive)/HR+ (hormone-receptor positive), and (4) HER2-positive (human epidermal growth factor receptor 2-positive)/HR- (hormone-receptor negative). Further details can be found elsewhere.[7]
7) *Nodal status*: Lymph node involvement was tagged by the presence of at least one ICD-10 diagnosis code for node disease (C773) between 250 days before and up to 180 days after early BC breast surgery. Nodal status was classified in a binary manner: "Node-positive" in presence of lymph node involvement; "Node-negative" otherwise.
8) *Type of BC surgery*: Breast surgery for BC was tagged with CCAM procedure codes at the hospital,[7] and were classified into two categories: (1) lumpectomy, and (2) mastectomy. The list of CCAM procedure codes can be found elsewhere.[7]
9) *Axillary surgery*: Axillar surgery for BC was tagged with CCAM procedure codes at the hospital,[7] and the use of axillar surgery was classified in a binary manner. The list of CCAM procedure codes can be found elsewhere.[7]
10) *Type of hospital for surgery:* The hospital where the patient received the initial breast surgery was categorized into (1) public hospital; (2) profit private hospital; (3) non-profit private hospital; and (4) CLCC ("*Centre de lutte contre le cancer"*, center for the fight against cancer). The information was directly available in the SNDS database.
11) U*se of radiotherapy*: Radiotherapy sessions were identified by ICD-10 diagnosis codes or CCAM procedure codes; and the use of radiotherapy was classified in a binary manner. The list of ICD-10 diagnosis codes and CCAM procedure codes can be found elsewhere.[7]

12) *Chemotherapy setting*: Chemotherapy sessions were identified by ICD-10 diagnosis codes, CCAM procedure codes and ATC molecule codes for hospital and outpatient care; and chemotherapy setting was classified into four categories: (1) no chemotherapy, (2) adjuvant chemotherapy only, (3) neoadjuvant chemotherapy only (NAC), and (4) both neoadjuvant and adjuvant chemotherapy. The list of ICD-10 diagnosis codes, CCAM procedure codes and ATC molecule codes can be found elsewhere.[7]

13) *Endocrine therapy regimen*: Endocrine therapy (ET) intake was flagged on the basis of outpatient delivery of (1) tamoxifen, (2) aromatase inhibitor (AI) or (3) gonadotropin-releasing hormone agonist (GnRH agonist). The type of ET delivered prior to trial inclusion was derived from a summary of all ET deliveries up to trial inclusion and was categorized into three groups: (1) no ET, (2) aromatase inhibitors (including aromatase inhibitor with or without GnRH agonists, GnRH agonists only, and switch from tamoxifen with or without GnRH agonists to aromatase inhibitors with or without GnRH agonists); (3) tamoxifen (including tamoxifen with or without GnRH agonists, and switch from aromatase inhibitors with or without GnRH agonists to tamoxifen with or without GnRH agonists). The list of outpatient delivery codes can be found elsewhere.[7]

14) *Adherence to endocrine therapy until trial inclusion:* The medication possession ratio (MPR) at trial inclusion for endocrine therapy was defined as the percentage of the number of endocrine therapy treatment days redeemed in outpatient care over the total number of days since endocrine therapy onset.[8] Adherence to endocrine therapy until trial inclusion was categorized into three groups: (1) Full adherence if the MPR was above 95%, (2) High adherence if the MPR was between 80% and 95%, and (3) Low adherence otherwise.

15) *Comorbid conditions in the year prior to inclusion*: The presence of a comorbid condition was detected on the basis of diagnosis and procedure codes in the year prior to inclusion, following the methodology described elsewhere.[9] Comorbid conditions were grouped into 12 categories: (1) cardiovascular, (2) endocrine and metabolism, (3) frailty, (4) gastrointestinal, (5) immune, (6) kidney, (7) liver, (8) neurologic, (9) psychiatric disorders, (10) pulmonary, (11) rheumatologic disease and connective tissue disorders, and (12) other.

16) *Exposure to other medications in the year prior to trial inclusion:* Medications redeemed the year prior to trial inclusion were identified from outpatient drug delivery data under prescription. Medications were classified according to the World Health Organization's Anatomical Therapeutic Chemical (ATC) classification; and were grouped into ATC second levels. A patient was considered to be exposed to a medication at trial inclusion if she received at least three months of the full dose in the six months prior to trial inclusion, following the methodology described elsewhere.[9] To avoid overparameterization of the model, only the ten most common ATC second levels were considered individually, so that exposure to other medications was categorized as: (1) drugs for acid-related disorders (ATC A02); (2) vitamins (ATC A11); (3) diuretics (ATC C03); (4) beta-blocking agents (ATC C07); (5) agents acting on the renin-angiotensin system (ATC C09); (6) lipid-modifying agents (ATC C10); (7) thyroid therapy (ATC H03); (8) analgesics (ATC N02); (9) psycholeptics (ATC N05); (10) psychoanaleptics (ATC N06); and (11) others.

17) *Age at trial inclusion:* Age at trial inclusion was calculated by the rounded difference, in years, between the date of trial inclusion and the date of birth.

#### Statistical analysis

The study population was described in terms of frequencies for qualitative variables and means and standard deviation for quantitative variables. Median follow-up and its interquartile range (IQR) were assessed by reverse Kaplan-Meier methods.

In the non-VET arm, where artificial censoring can occur any month during the grace period, we derived IPC weights from a Cox proportional hazards model regressing artificial censoring on all covariates. IPC weights were updated monthly during the grace period (12 months) and then remained fixed until either administrative censoring or the event of interest. In the VET arm, where artificial censoring can only occur at the end of the grace period, we derived IPC weights from a logistic regression model fitting artificial censoring on all covariates. IPC weights were fixed to 1 during the grace period (12 months), were updated once at the end of the grace period, and then remained fixed until either administrative censoring or the event of interest.[1]

ATE was estimated using the three-year and five-year differences in DFS on the IPC weighted dataset. In contrast to Cox hazard ratios, survival differences have a straightforward causal interpretation regardless of whether the proportional hazards assumption holds.[11] We computed bootstrap confidence intervals with 1,000 iterations on a smaller dataset consisting of all patient-years initiating VET and 20% of patient-years non initiating VET to generate 95% confidence intervals. In the SNDS, which covers all health care consumption in France, including the Overseas Departments, loss to follow-up was only possible due to emigration. We did not take this further into account in the analysis.

In a sensitivity analysis, we used fosfomycin initiation as a negative control exposure, with DFS as outcome. We used the same targeted trial emulation strategy with identical inclusion/exclusion criteria, except that absence of VET delivery for at least two years prior to trial inclusion became absence of fosfomycin delivery. Patients were classified as initiating fosfomycin if they filled at least two prescriptions for fosfomycin (identified by ATC code J01XX01) in the year following inclusion in the emulated trial. As with the VET analysis, patients with a single delivery during the grace period were discarded from both arms during the grace period. The statistical estimation of the ATE for the initiation of fosfomycin was identical to the one used in the VET analysis.

In a supplementary analysis aiming at mimicking an associational methodology previously proposed,[12] we fitted a time-dependent Cox proportional hazards regression model. The model was fitted on all women, diagnosed with a non-metastatic BC treated at least by breast surgery and any systemic treatment (chemotherapy, targeted therapy, or endocrine therapy), and aged between 40 and 79 years old at BC diagnosis. Patients who relapsed or died in the first six months after cancer diagnosis were excluded. VET initiation was modelled as a time-varying variable, with a lag of six months. Covariates included in the model were age (continuous), year of BC diagnosis (continuous), deprivation level quintile of the area of residence, type of BC surgery (lumpectomy/mastectomy), chemotherapy (binary), radiotherapy (binary), nodal status, comorbid conditions at the time of BC diagnosis, other medication use at the time of BC diagnosis (including statins, aspirin, metformin, and any systemic use of sexual hormones), use of tamoxifen (time-varying, with a lag of six months), and use of aromatase inhibitors (time-varying, with a lag of six months). The outcome was time to BC recurrence or death. Subgroup analyses were conducted based on endocrine therapy regimen, categorized into (i) no tamoxifen and no AI at any time during follow-up, (ii) tamoxifen only, started at any time during follow-up, and (iii) AI with or without tamoxifen, started at any time during follow-up. Of note: the subgroups were defined using post-inclusion information, as it was done in the previous study, although this procedure may induce bias. An extensive comparison between the inclusion criteria, exposure definition, covariates, and outcome proposed in McVicker et al.[12] and our analysis is presented in Supplementary Table 3.

Analyses were performed with R software, version 3.6.3. Logistic regression models were computed with the *glm* function from the R base package stats. Standardized mean differences (SMDs) were computed with the *bal.tab* function from the R package cobalt (version 4.1.0). Cox proportional hazards models were fitted with the *coxph* function from the R package survival (version 3.2-3). Survival curves were drawn with the R package survminer (version 0.4.7). All confidence intervals were two-sided.

#### Supplementary Tables

| Commercial name | Estrogen Molecule | Presentation |
|---|---|---|
| *Physiogine©* | Estriol | Ovule |
| *Trophigil©** | Estriol | Ovule |
| *Florygynal©** | Estriol | Ovule |
| *Colpotrophine©* | Promestriene | Ovule |
| *Physiogine© 0,1 %* | Estriol | Cream |
| *Gydrelle© 0,1%* | Estriol | Cream |
| *Trophicreme© 0,1%* | Estriol | Cream |
| *Colpotrophine© 1%* | Promestriene | Cream |

**Supplementary Table 1: List of medications considered as vaginal estrogen treatment, with molecule (promestriene or estriol) and presentation (cream or ovule).**

*: In combination with progesterone.

| | | Medical data | CCCAM | ICD-10 | ATC |
|---|---|---|---|---|---|
| (i) Treatment resumption at least 6 months after the end of the initial treatments | Radiotherapy | Appointment for antineoplastic radiation therapy | | Z5100 | |
| | | Radiotherapy preparation session | | Z5101 | |
| | | Radiotherapy session | | Z510 | |
| | | Radiotherapy procedures | ZZNA002, ZZNL001, ZZNL002, ZZNL003, ZZNL004, ZZNL005, ZZNL006, ZZNL009, ZZNL011, ZZNL012, ZZNL013, ZZNL014, ZZNL015, ZZNL016, ZZNL017, ZZNL018, ZZNL019, ZZNL02, ZZNL030, ZZNL031, ZZNL036, ZZNL037, ZZNL039, ZZNL040, ZZNL042, ZZNL043, ZZNL045, ZZNL046, ZZNL048, ZZNL049, ZZNL050, ZZNL051, ZZNL052, ZZNL053, ZZNL054, ZZNL055, ZZNL058, ZZNL059, ZZNL060, ZZNL061, ZZNL062, ZZNL063, ZZNL064, ZZNL065, ZZNL900, ZZNL902, ZZNL903, ZZNL904, ZZNL905, ZZNL906, YYYY016, YYYY021, YYYY023, YYYY045, YYYY046, YYYY047, YYYY048, YYYY049, YYYY050, YYYY051, YYYY052, YYYY053, YYYY054, YYYY055, YYYY056, YYYY080, YYYY081, YYYY099, YYYY101, YYYY109, YYYY122, YYYY128, YYYY136, YYYY141, YYYY151, YYYY152, YYYY166, YYYY175, YYYY197, YYYY1211, YYYY223, YYYY225, YYYY244, YYYY256, YYYY267, YYYY299, YYYY301, YYYY302, YYYY303, YYYY304, YYYY305, YYYY306, YYYY307, YYYY310, YYYY312, YYYY313, YYYY314, YYYY315, YYYY316, YYYY320, YYYY323, YYYY324, YYYY325, YYYY326, YYYY327, YYYY331, YYYY334, YYYY335, YYYY336, YYYY337, YYYY338, YYYY343, YYYY345, YYYY346, YYYY347, YYYY348, YYYY349, YYYY356, YYYY357, YYYY358, YYYY359, YYYY360, YYYY365, YYYY367, YYYY368, YYYY369, YYYY370, YYYY371, YYYY377, YYYY379, YYYY380, YYYY381, YYYY382, YYYY383, YYYY387, YYYY390, YYYY391, YYYY392, YYYY393, YYYY398, YYYY450, YYYY451, YYYY457, YYYY458, YYYY459, YYYY460, YYYY468, YYYY469, YYYY470, YYYY471, YYYY479, YYYY480, YYYY481, YYYY491, YYYY492, YYYY493, YYYY497, YYYY500, YYYY511, YYYY520, YYYY522, YYYY533, YYYY544, YYYY555, YYYY566, YYYY577, YYYY588, YYYY599 | | |
| | Chemotherapy | Encounter for antineoplastic chemotherapy and immunotherapy | | Z511, Z512 | |
| | | Anticancerous agent administration | ZZLF900 | | |
| | | Port implanting | EBLA001 | | |
| | | Cerebral anticancerous agent administration | ABLB006 | | |
| | | Intrathecal anticancerous agent administration | AFLB003, AFLB013 | | |
| | | Upper limb arterial anticancerous agent administration | ECLF005, ECLF006 | | |
| | | Lower limb arterial anticancerous agent administration | EELF004, EELF005 | | |
| | | Liver anticancerous agent *in situ* administration | EDLF014, EDLF015, EDLF016, EDLF017 | | |
| | | Kidney anticancerous agent *in situ* administration | EDLF018, EDLF019, EDLF020, EDLF021 | | |
| | | Cervical/cephalic anticancerous agent administration | EBLF002, EBLF003 | | |
| | | Intrapleural anticancerous agent administration | GGLB001, GGLB008 | | |
| | | Intraperitoneal anticancerous agent administration | HPLB002, HPLB003, HPLB007 | | |
| | | Locoregional anticancerous agent administration with extracorporeal circulation | ZZLF004 | | |
| | | Locoregional anticancerous agent administration without extracorporeal circulation | ZZLF900 | | |
| | | CYCLOPHOSPHAMIDE | | | L01AA01 |
| | | DOCETAXEL (identifiable until March 2012) | | | L01CD02 |
| | | EPIRUBICIN | | | L01DB03 |
| | | FLUOROURACIL | | | L01BC02 |
| | | PACLITAXEL | | | L01CD01 |
| | Targeted Therapy | PERTUZUMAB | | | L01XC13 |
| | | TRASTUZUMAB | | | L01XC03, L01XC14 |
| (ii) Breast surgery procedure with axillar procedure performed at least 6 months after the end of the initial treatments | | Mastectomy with axillary surgery | QEFA003, QEF005, QEFA010, QEFA020 | | |
| | | Partial mastectomy with axillary surgery | QEFA001, QEFA008 | | |
| | | Mastectomy without axillary surgery in combination with axillary surgery without breast surgery | QEFA007, QEFA012, QEFA013, QEFA015, QEFA019 in combination with FCFA021, FCFA029 | | |
| | | Partial mastectomy without axillary surgery in combination with axillary surgery without breast surgery | QEFA004, QEFA016, QEFA017, QEFA018 in combination with FCFA021, FCFA029 | | |
| (iii) Intake of an anti-cancer molecule approved only in the metastatic setting starting at least six months after initial breast surgery | Chemotherapy molecule | CAPECITABINE | | | L01BC06 |
| | | CARBOPLATIN | | | L01XA02 |
| | | CISPLATIN | | | L01XA01 |
| | | DOXORUBICIN (only CAELYX®) | | | L01DB01 |
| | | ERIBULIN | | | L01XX41 |
| | | ETOPOSIDE | | | L01CB01 |
| | | GEMCITABINE | | | L01BC05 |
| | | MELPHALAN | | | L01AA03 |
| | | METHOTREXATE | | | L01BA01 |
| | | MILTEFOSINE | | | L01XX09 |
| | | MITOMYCIN | | | L01DC03 |
| | | MITOXANTRONE | | | L01DB07 |
| | | PIRARUBICIN | | | L01DB08 |
| | | THIOTEPA | | | L01AC01 |
| | | VINBLASTINE | | | L01CA01 |
| | | VINCRISTINE | | | L01CA02 |
| | | VINDESINE | | | L01CA03 |
| | | VINORELBINE | | | L01CA04 |
| | *HER2*- Targeted Therapy *molecule* | LAPATINIB | | | L01XE07 |
| | Non-*HER2* Targeted Therapy molecule | BEVACIZUMAB | | | L01XC07 |
| | | BYL719 (ALPELISIB) | | | L01XE |
| | | EVEROLIMUS | | | L01XE10 |
| | | PALBOCICLIB | | | L01XE33 |
| | Endocrine Therapy molecule | TOREMIFENE | | | L02BA02 |
| | | FORMESTANE | | | L02BG02 |
| | | FULVESTRANT | | | L02BA03 |
| (iv) Presence of a diagnosis code of metastasis in hospitalization stays starting at least six months after initial breast surgery | | Metastatic disease | | C77 except C773, C78, C79 | |

**Supplementary Table 2: List of CCAM, ICD-10 and ATC codes used to identify breast cancer relapses.** Abbreviations: ICD 10: international statistical classification and related health problems – 10th revision; CCAM: "Classification Communes des Actes Médicaux"; ATC: anatomical therapeutic and chemical classification

| | | | *McVicker et al.* | *Dumas et al. - supplementary analysis* |
|---|---|---|---|---|
| **Method** | **Inclusion criteria** | | Women, aged 40 to 79, newly diagnosed with early breast cancer between 2010 to 2017 in Scotland and 2000 to 2016 in Wales. Patients previously diagnosed with other invasive cancer diagnoses were excluded. Patients who died in the first 6 months after cancer diagnosis were excluded. | Women, aged 40 to 79, newly diagnosed with a non-metastatic breast cancer between 2011 and 2017 in France, treated at least by breast surgery and any systemic treatment (chemotherapy, targeted therapy, or endocrine therapy). Patients with concomitant cancer of another site were excluded. Patients who relapsed or died in the first 6 months after cancer diagnosis were excluded. |
| | **Exposure** | | Exposure was modelled as a single time-varying variable, with a lag of 6 months, into the following hierarchical categories: systemic HRT use (with or without vaginal estrogen), vaginal estrogen therapy only use and HRT non-use. HRT use was ascertained from electronic general practitioner (GP) prescribing records (Wales) or dispensing records (Scotland). | Exposure was modelled as a single time-varying variable, with a lag of 6 months, into VET use or non VET use. VET use was ascertained from dispensing records. |
| | **Outcome** | | Time to breast cancer–specific mortality | Time to disease recurrence or death |
| | **Covariates** | | Age, year, deprivation level, cancer treatment (surgery, chemotherapy, and radiotherapy), tamoxifen or aromatase inhibitor use (as time-varying covariates with 6-month lags), Charlson Comorbidity Index (before diagnosis), anemia (before diagnosis), other medication use (including statins, aspirin, metformin, and oral contraceptives before diagnosis), hysterectomy or oophorectomy (anytime up to 6 months after diagnosis), and cancer stage and grade. Where missing, cancer stage and grade were imputed using multiple imputation with chained equations. | Age, year, deprivation level, cancer treatment (type of surgery, chemotherapy, and radiotherapy), tamoxifen or aromatase inhibitor use (as time-varying covariates with 6-month lags), comorbid conditions before diagnosis, other medication use before diagnosis (including statins, aspirin, metformin, and any systemic use of sexual hormones), and nodal status. There was no missing data. |
| | **Model** | | Time-dependent proportional hazards Cox model | Same |
| **Results** | **Number of patients** | | 49 237 | 165 993 |
| | **Number of patient-years** | | 300 673 | 338 666 |
| | **Number of patient-years initiating VET** | | 11 437 | 18 747 |
| | **Number of events in the VET group** | | 120 | 1 263 |
| | **HR (95% CI)** | Whole population | 0·77 (0·63-0·94) | 0·95 (0·89-1·00) |
| | | *No tamoxifen and no AI* | *0·67 (0·43-1·04)* | *0·99 (0·87-1·12)* |
| | | *Tamoxifen only* | *1·01 (0·52-1·95)* | *0·96 (0·82-1·12)* |
| | | *AI with or without tamoxifen* | *0·72 (0·58-0·91)* | *0·93 (0·87 -1·00)* |

**Supplementary Table 3: Summary and comparison of methods and results for the supplementary analysis mimicking an associational method previously proposed to investigate the effect of VET use on oncological outcomes.** Abbreviations: HR: hazard ratio, CI: confidence interval, VET: Vaginal estrogen therapy, HRT: hormone replacement therapy; AI: aromatase inhibitors.

| | VET initation | | | No VET initation |
|---|---|---|---|---|
| | *Any VET* | *Promestriene only* | *Estriol only* | |
| *Number of patient-years included* | 1,739 (0·5%) | 965 (0·3%) | 601 (0·2%) | 366,858 (98·2%) |
| *Number of events** | 156 (9·0%) | 95 (9·8%) | 45 (7·5%) | 31,668 (8·6%) |

**Supplementary Table 4: Number of patient-years initiating VET, patient-years initiating promestriene, patient-years initiating estriol, patient-years non initiating any VET, and outcome events for DFS for the pooled analysis of all sequential trials.**

Of note: of the 373,568 patient-years meeting the inclusion criteria, 4,971 patient-years received VET only once during the grace period and were artificially censored from both arms. This explains why the number of patient-years of the "any VET" and in the "no VET initiation" groups do not sum to 373,568, and why the percentages do not sum to 100. * The percentage is calculated in relation to the line above. Abbreviations : VET: vaginal estrogen therapy; DFS: disease-free survival.

| | | No VET | Any VET |
|---|---|---|---|
| ***Number of patient-years*** | | **366858** | **1739** |
| **Comorbid conditions** | | | |
| Psychiatric | | 32684 (8·9) | 159 (9·1) |
| Rheumatologic and connectivopathies | | 7236 (2·0) | 38 (2·2) |
| Pulmonary | | 9317 (2·5) | 50 (2·9) |
| Endocrine and metabolism | | 45015 (12·3) | 218 (12·5) |
| Cardiovascular | | 39621 (10·8) | 193 (11·1) |
| Frailty | | 19529 (5·3) | 99 (5·7) |
| Gastrointestinal | | 7743 (2·1) | 59 (3·4) |
| Any other comorbid condition | | 11944 (3·3) | 40 (2·3) |
| **Exposure to other medications** | | | |
| Drugs for acid related disorders (ATC A02) | | 47252 (12·9) | 346 (19·9) |
| Vitamins (ATC A11) | | 140598 (38·3) | 849 (48·8) |
| Diuretics (ATC C03) | | 38982 (10·6) | 170 (9·8) |
| Beta blocking agents (ATC C07) | | 38390 (10·5) | 185 (10·6) |
| Agents acting on the renin-angiotensin system (ATC C09) | | 57571 (15·7) | 240 (13·8) |
| Lipid modifying agents (ATC C10) | | 48404 (13·2) | 272 (15·6) |
| Thyroid therapy (ATC H03) | | 43463 (11·8) | 229 (13·2) |
| Analgesics (ATC N02) | | 103667 (28·3) | 600 (34·5) |
| Psycholeptics (ATC N05) | | 59216 (16·1) | 364 (20·9) |
| Psychoanaleptics (ATC N06) | | 48798 (13·3) | 307 (17·7) |
| Any other medication | | 182123 (49·6) | 1014 (58·3) |
| **Endocrine therapy (among *HR* -positive tumors only)** | | | |
| Endocrine therapy regimen in the last year | Aromatase inhibitors (AIs) | 181469 (57·9) | 972 (65·0) |
| | tamoxifen | 129962 (41·5) | 510 (34·1) |
| | No ET | 2080 (0·7) | 14 (0·9) |
| ET adherence | Full adherence | 228252 (72·8) | 1158 (77·4) |
| | High adherence | 37087 (11·8) | 149 (10·0) |
| | Low adherence | 42293 (13·5) | 156 (10·4) |
| | Unknown* | 5879 (1·9) | 33 (2·2) |

**Supplementary Table 5: Patient's characteristics by VET initiation status for all variables repeatedly measured, at the time of each target trial inclusion.** The number of patients-years, and the percentage of patients-years (in parentheses), are reported for categorical variables. *ET adherence was unknown if the patient received only gonadotropin-releasing hormone (gnRH) agonists or did not start ET prior to the start of the emulated trial. Patients receiving only gnRH agonists were included in the aromatase inhibitor group for the endocrine therapy regimen. Of note: of the 373,568 patient-years meeting the inclusion criteria, 4,971 patient-years received VET only once during the grace period and were artificially censored from both arms. They are not shown in this table. *Abbreviations*: VET: vaginal estrogen therapy.

| | Fosfomycin initiation | No fosfomycin initation |
|---|---|---|
| *Number of patient-years included* | 2,959 (0·8%) | 334,675 (95·3%) |
| *Number of events** | 225 (7·6%) | 29,502 (8·8%) |

**Supplementary Table 6: Number of participants (in patient-years), patient-years initiating fosfomycin, patient-years non initiating fosfomycin and outcome events for DFS for the pooled analysis of all sequential trials for the negative control exposure analysis.**

Of note: of the 351,128 patient-years meeting the inclusion criteria, 13,494 patient-years received fosfomycin only once during the grace period and were artificially censored from both arms. This explains why the number of patient-years of the "Fosfomycin initiation" and in the "no fosfomycin initiation" groups do not sum to 351,128, and why the percentages do not sum to 100 in the first row. * The percentage is calculated in relation to the line above. Abbreviations: VET: vaginal estrogen therapy; DFS: disease-free survival.

| | **Whole population** | ***HR*- positive** | | | ***HR* -negative** |
|---|---|---|---|---|---|
| | | All | *Currently treated with AI* | *Currently treated with tamoxifen* | |
| **3 years** | 0·1 (-0·9 to 1) | -0·3 (-1·3 to 0·9) | -0·9 (-2·3 to 0·8) | 0·4 (-1·2 to 2) | 2·2 (-0·9 to 4·3) |
| **5 years** | 0 (-1·7 to 1·9) | -0·8 (-2·5 to 1·4) | 0·1 (-1·7 to 3) | -1·3 (-4·8 to 1·8) | 4·7 (0·8 to 7·8) |

**Supplementary Table 7: Difference in disease-free survival (DFS, in percentage-points) at three years and five years and associated 95% confidence intervals for fosfomycin initiation versus no initiation for the whole population and per subgroup.**

Abbreviations: HR: hormone receptor; AI: aromatase inhibitor.

| First Author | Study title | Year | Journal | Country | Study Design | Number of patients | Number of VET users | Patient charateristics | Endocrine therapy | Type of VET | Definition of VET users | Outcome | Median time (range) of follow up | Results (VET versus no VET) |
|---|---|---|---|---|---|---|---|---|---|---|---|---|---|---|
| **Dumas** | *Impact of vaginal estrogen therapy initiation after breast cancer on oncological outcomes: emulation of nationwide population-based target trials* | 2024 | | France | Retrospective cohort<br>Emulated target trial | 134942 | 1737 | Stade I-III | TAM<br>AI<br>None | Promestriene: 55·5%<br>Estriol: 34·6%<br>Promestriene and estriol: 9·9% | At least 2 prescriptions | BC recurrence | 33 months (16-50) | • BC recurrence at 3 years: 0 percentage-point (95% CI: -0·9 to 1·5)<br>• BC recurrence at 5 years: -1·6 percentage-point (95% CI: -4·0 to 0·3)<br>In patients currently treated with AI:<br>• BC recurrence at 3 years: - 0·6 percentage-point (95% CI: -2·2 to 1·2)<br>• BC recurrence at 5 years: -3·0 percentage-point (95% CI: -6·5 to -0·3)<br>In patients currently treated with tamoxifen:<br>• BC recurrence at 3 years: 2·2 percentage-point (95% CI: 0·2 to 4)<br>• BC recurrence at 5 years: 0·0 percentage-point (95% CI: -4·5 to 3·9) |
| **McVicker** | *Vaginal Estrogen Therapy Use and Survival in Females With Breast Cancer* | 2023 | Jama Oncol | Scotland and Wales | Retrospective cohort | 49 237 | 2551 | Stade I-IV | TAM<br>AI<br>None | Estriol<br>Estradiol | At least 1 prescription | BC mortality | 8 years (5-12) (Wales cohort)<br>5 years (3-7) Scotland cohort ) | • BC mortality: HR= 0·77 (95%CI: 0·63-0·94)<br>• BC mortality for women using AI, with or without tamoxifen: HR= 0·99 (95%CI: 0·79-1·24)<br>• BC mortality for women using tamoxifen only: HR= 0·41 (95%CI: 0·21-0·79) |
| **Agrawal** | *Safety of Vaginal Estrogen Therapy for Genitourinary Syndrome of Menopause in Women With a History of Breast Cancer* | 2023 | Obstetrics and gynecology | USA | Retrospective cohort | 42113 | 2111 | Stade I-III<br>GSM occuring 3 months to 5 years after BC diagnosis | AI<br>None | Estradiol<br>Conjugated equine estrogen | Three or more vaginal estrogen , with at least one prescription within 1 year after their initial GSM diagnosis | BC recurrence | 5 and 10 years | • BC recurrence: HR= 0·98 (95%CI: 0·85–1·14)<br>• Overall mortality at 5 years: RR= 0·74 (95%CI: 0·53–1·03)<br>• Overall mortality at 10 y: RR= 0·70 (95%CI: 0·51–0·96)<br>In HR+ BC:<br>• BC recurrence: HR= 0·88 ( 95%CI: 0·69–1·12)<br>In VET group:<br>• Risk of BC recurrence (AI users vers AI no users): RR= 5 (95%CI: 3·05–8·19) |
| **Sund** | *Estrogen therapy after breast cancer diagnosis and breast cancer mortality risk* | 2023 | Breast Cancer Research and Treatment | Sweden | Retrospective Case control | 15 198 | 2359* | Stade I-III<br>HR-positive breast cancer<br>Treatment with endocrine therapies | TAM<br>AI | Estriol<br>Estradiol | At least 1 prescription | BC mortality | 5·1 years (0·4-13·3) | • BC mortality for patients treated by AI: OR=0·87 (95%CI: 0·62–1·22)<br>• BC mortality for patients treated by tamoxifen: OR=1·30 (95%CI: 0·91–1·86) |
| **Cold** | *Systemic or Vaginal Hormone Therapy After Early Breast Cancer: A Danish Observational Cohort Study* | 2022 | *Journal of the National Cancer Institute* | Denmark | Retrospective cohort | 8461 | 1957 | Stade I-III<br>Post menopausal women<br>HR-positive BC<br>No chemotherapy | AI: 4·8%<br>AI/TAM: 34·7%<br>TAM: 23·7%<br>None: 36·8% | Estradiol<br>Estriol | At least 2 prescriptions | BC recurrence<br>Overall mortality | 9·8 years for recurrence<br>15·2 years for death | · BC recurrence: HR= 1·08 (95%CI: 0·89 -1·32)<br>Subgroup receiving AI or sequential AI/Tamoxifen : HR=1·39 (95%CI: 1·04 -1·85)<br>· Total death : HR= 0·78 (95%CI: 0·71-0·87) |
| **Le Ray** | *Local estrogen therapy and risk of breast cancer recurrence among hormone-treated patients: a nested case-control study* | 2012 | *Breast Cancer Research and Treatment* | UK | Retrospective Case control | 13479 | 271 | Stade I-III<br>Treated by TAM or AI | TAM: 84%<br>AI: 16% | Estrogens (no detail) | ND | BC recurrence | 4·2 years | • BC recurrence for women treated by TAM or AI: RR= 0·78 (95 %CI: 0·48–1·25)<br>Stratified analysis:<br>· TAM : RR= 0·83 (95%CI 0·51–1·34)<br>· AI: to few patients |
| **Dew** | *A cohort study of topical vaginal estrogen therapy in women previously treated for breast cancer* | 2003 | *Climateric* | Australia | Retrospective cohort | 1472 | 69 | Stade I-IV<br>Post menopausal women | TAM (47%)<br>None (53%) | Estriol: 52%<br>Estradiol: 48% | At least 1 prescription | BC recurrence | 5·5 years (0·5-29) | · BC recurrence: HR= 0·57 (95%CI: 0·20-1·58)<br>· BC mortality: to few death registered and adjusted analysis not possible |
| **Durna** | *Hormone replacement therapy after a diagnosis of breast cancer: cancer recurrence and mortality* | 2002 | *The Medical Journal of Australia* | Australia | Retrospective cohort | 1122 | 32 | Stade I-IV<br>Post menopausal women | TAM (60%)<br>None | Estriol<br>Estradiol | ND | BC recurrence<br>BC mortality<br>Overall mortality | 6·08 years | · BC recurrence : RR= 0·18 (95%CI: 0·04–0·75)<br>· BC mortality : RR= 0·35 (95%CI: 0·07–1·68)<br>· Overall mortality : RR= 0·30 (95%CI: 0·07–1·30) |
| **O'Meara** | *Hormone Replacement Therapy After a Diagnosis of Breast Cancer in Relation to Recurrence and Mortality* | 2001 | *Journal of the National Cancer Institute* | USA | Retrospective cohort | 869 | 75 | Stage I-III | TAM (21·9%)<br>None (78·1%) | Conjugated equine estrogen<br>Dienestrol | At least 2 prescriptions within a 6-month interval | BC recurrence<br>BC mortality<br>Overall mortality | 5·8 years (0–29) | · BC recurrence: RR= 0·46 (95%CI: 0,21-1,01)<br>· BC mortality RR= 0·37 (95%CI: 0·11-1·21)<br>· Overall mortality: RR= 0·60 (95%CI: 0·31-1·16) |
| **Vassilopoulou-Sellin** | *Estrogen Replacement Therapy in Women with Prior Diagnosis and Treatment for Breast Cancer* | 1997 | *Gynecologic Oncology* | USA | Prospective | 6 | 6 | Stage I-III | ND | Conjugated equine estrogen | Received VET for a minimun of 2 years | BC recurrence | 95 months | No cancer recurrences in patient group |

**Supplementary Table 8: Summary of previous studies investigating the impact of VET use on oncologic outcomes for patients with previous BC.**

*Authors were not able to distinguish the pharmaceutical form of estrogen therapy (VET or HRT) but they presume that the vast majority of estrogen therapy prescribed in their study was VET. Abbreviations: AI: aromatase inhibitor; TAM: tamoxifen; ND: Not done; BC: Breast cancer; VET: vaginal estrogen therapy; HRT: hormone replacement therapy.

#### Supplementary Figures

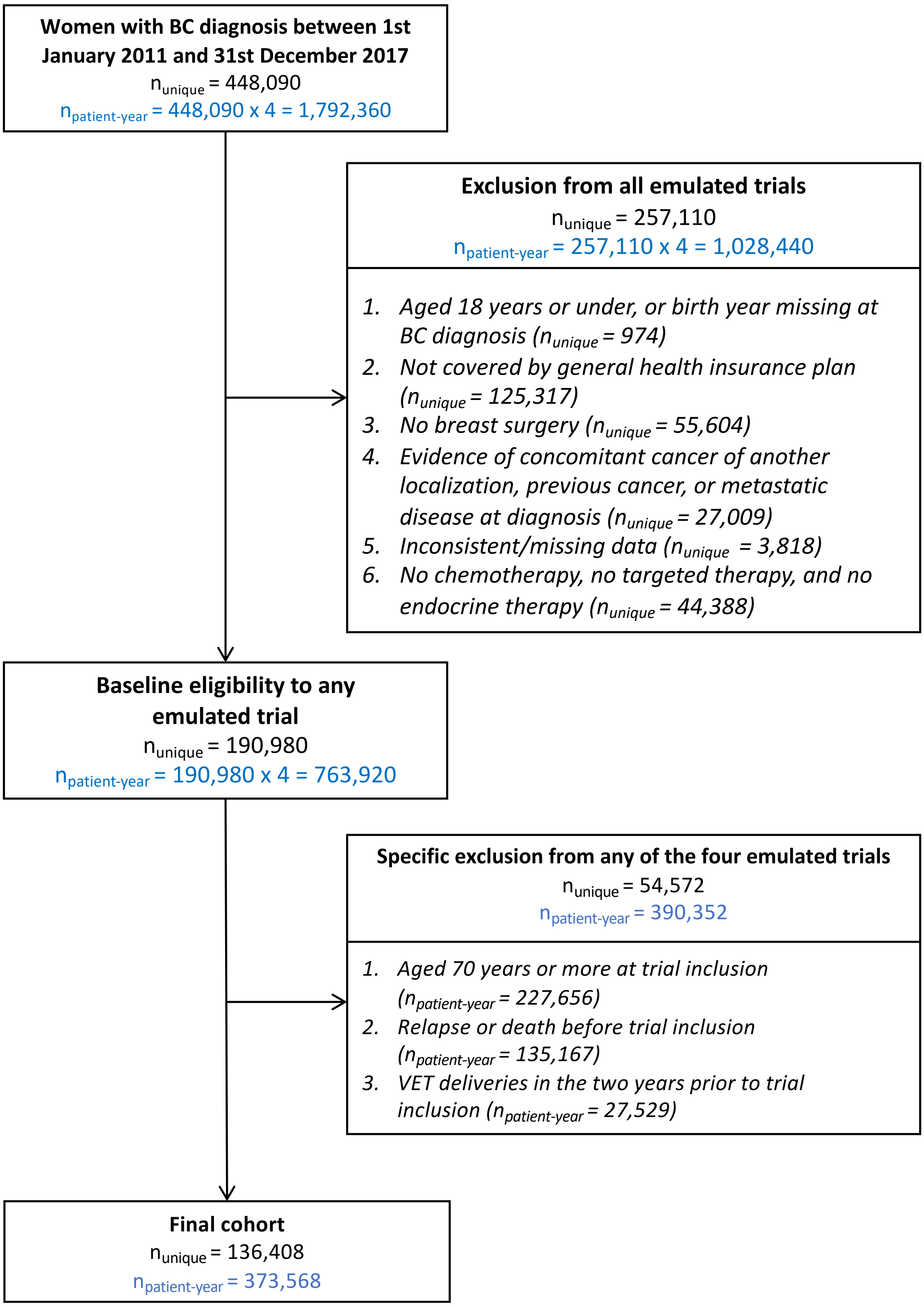

**Supplementary Figure 1: Study flowchart.**

*Abbreviations: BC:* breast cancer; VET: vaginal estrogen therapy.

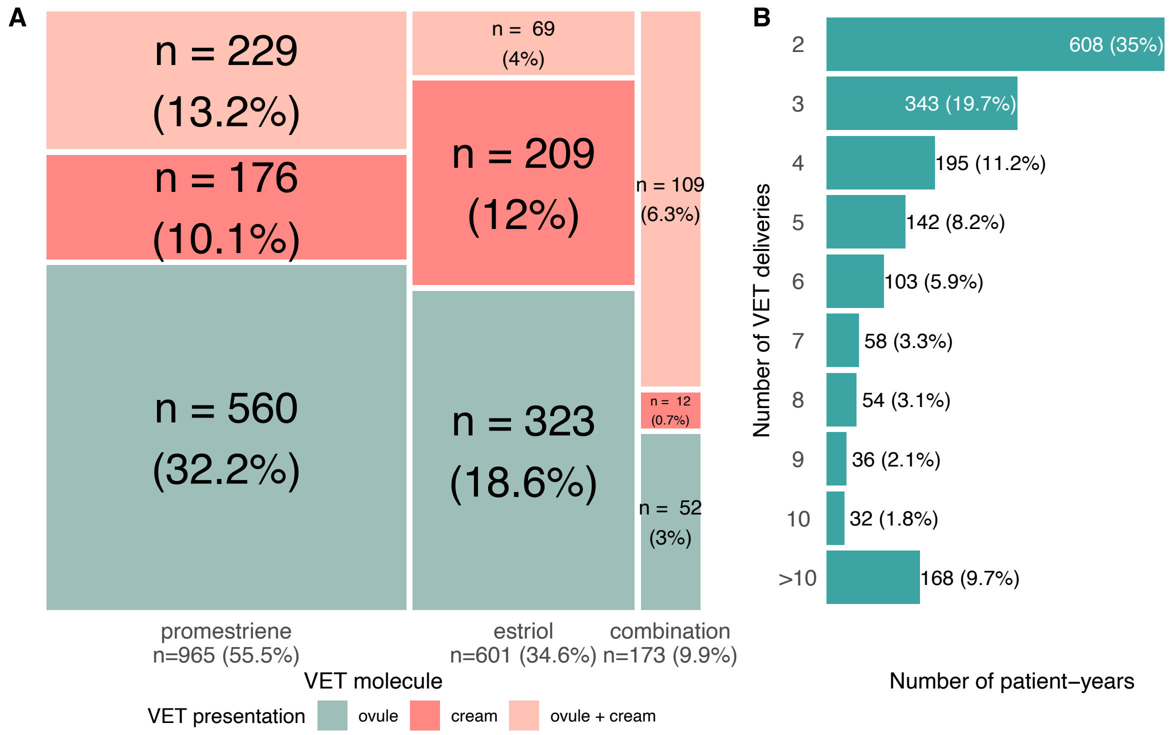

**Supplementary Figure 2: Distribution of VET molecule, presentation and number of deliveries**

(A) Distribution of VET molecule (promestriene, estriol, or combination) and presentation (ovule, cream, or both) in patients initiating VET for the first two deliveries. (B) Distribution of number of VET deliveries in patients initiating VET. *Abbreviations: VET:* Vaginal Estrogen Therapy

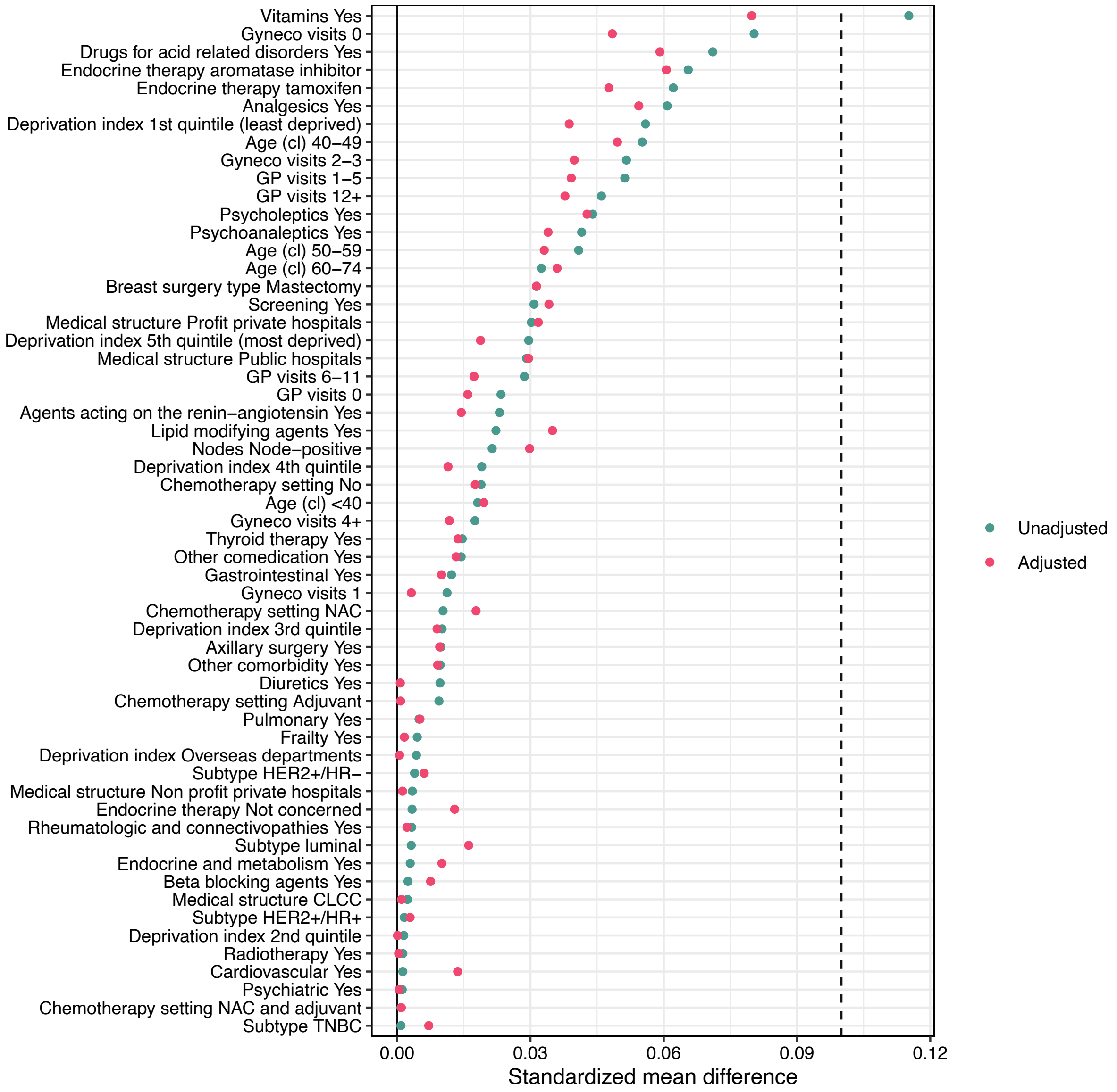

**Supplementary Figure 3: Standardized mean differences (SMDs) before (unadjusted) and after (adjusted) inverse probability of censoring weighting for disease-free survival analysis for all covariates.**

Covariates are ordered by decreasing SMD before weighting. *Abbreviations:* GP: general practitioner; NAC: neoadjuvant chemotherapy; HER2: human epidermal growth factor receptor 2; HR: hormone receptor; CLCC: TNBC: triple negative breast cancer; CLCC: “Centre de Lutte Contre le Cancer” (center for the fight against cancer).

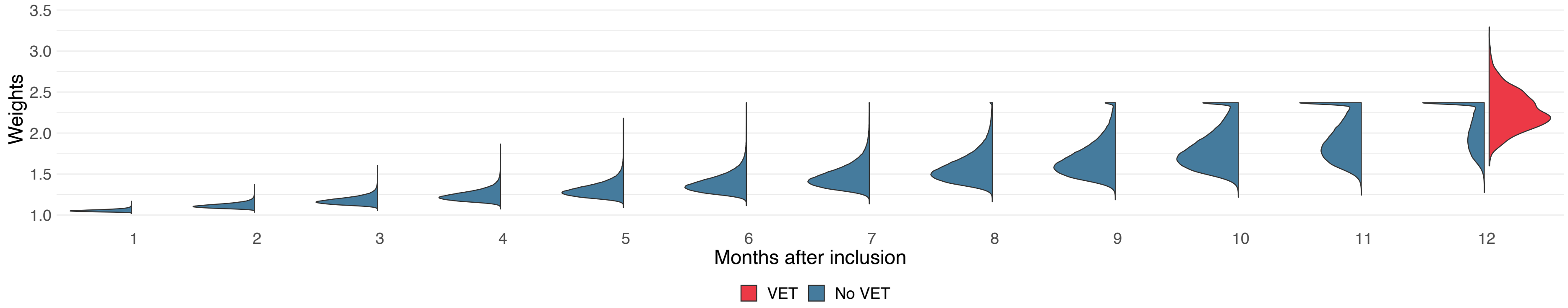

**Supplementary Figure 4: Distribution of time-varying inverse probability of censoring weights by months during the grace period from the VET (treatment) and no VET (control) arm.**

Abbreviations: VET: vaginal estrogen therapy.

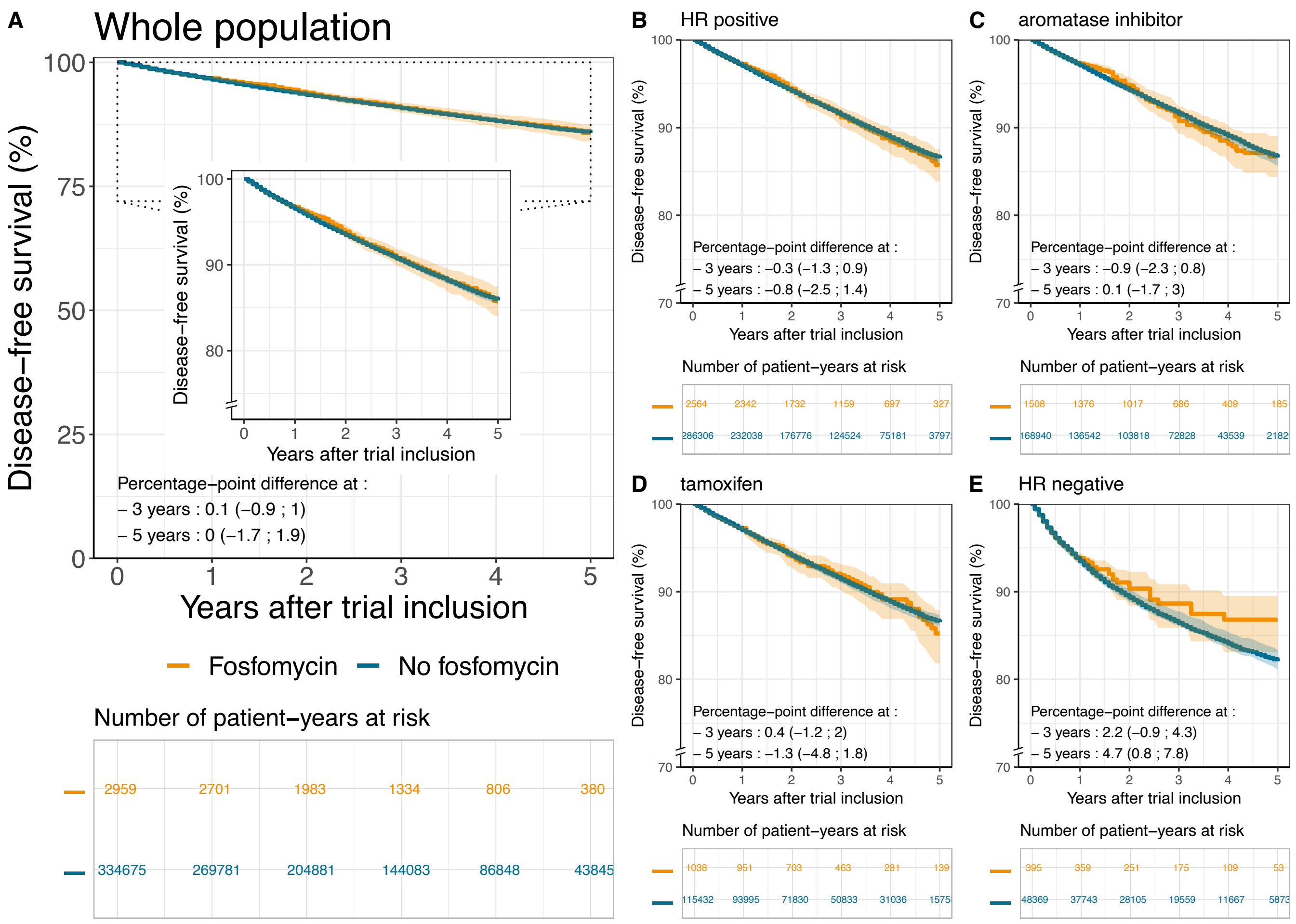

**Supplementary Figure 5: Effects of fosfomycin initiation on disease-free survival for the whole population of BC patients (A) and for subgroups defined by HR status and endocrine therapy regimen (B-E).**

Abbreviations: HR: hormone receptor; CI: confidence interval.
